# Mapping inequalities in dementia diagnosis in England: a Bayesian spatial analysis of disease prevalence and case ascertainment

**DOI:** 10.64898/2026.11.09.26367595

**Authors:** Beatrice Taylor, Jessica Jiang, Jason D. Warren, Sebastian J. Crutch

## Abstract

**Background:** By 2030, over one million people in the UK are projected to be living with dementia; however, the NHS estimates that only approximately 65% of individuals with dementia currently receive a formal diagnosis. Understanding the geographic and socioeconomic factors contributing to variation in diagnosis is essential for reducing inequalities in dementia care.

**Methods:** We developed a Bayesian spatial modelling framework to separate underlying dementia prevalence from case ascertainment using routinely collected public datasets at Local Authority District (LAD) level across England. Underlying prevalence was modelled using non-modifiable and modifiable risk factors identified by the Lancet Commission, while diagnosis rates were modelled as a function of healthcare provision and sociodemographic factors. Spatial random effects were incorporated to identify residual geographic variation after accounting for observed covariates. We additionally compared case ascertainment with regional variation in dementia subtype coding as a further measure of diagnostic quality.

**Findings:** Substantial geographic variation in dementia diagnosis remained after accounting for differences in underlying disease prevalence. Urbanicity, deprivation, ethnicity, and healthcare provision were associated with variation in case ascertainment, with spatial effects explaining additional regional variation. Diagnostic specificity varied considerably between regions, with areas of lower case ascertainment tending to exhibit higher proportions of undifferentiated dementia diagnoses.

**Interpretation:** By jointly modelling underlying dementia prevalence and case ascertainment, we can distinguish the factors associated with dementia burden from those associated with diagnosis. This enables the identification of areas with low case ascertainment, which when combined with poor diagnostic specificity, provides an evidence base for targeted regional interventions. Potential interventions include strengthening dementia care pathways through enhanced outreach, more equitable access to diagnostic assessment, and locally tailored models of care.

## 1 Introduction

Dementia prevalence is influenced by both non-modifiable and modifiable risk factors. The main non-modifiable risk factors are age, sex [1] and genetic susceptibility [2], of which the most significant is age with the incidence of all-cause dementia increasing markedly in later life (reaching 18.2% per year among the oldest age groups) [3]. The most recent Lancet commission on dementia [4] identified a further 14 modifiable risk factors, defined as potentially reversible lifestyle factors. In early life this is: less education; in mid-life: hearing loss, high cholesterol, depression, traumatic brain injury, physical inactivity, diabetes, smoking, hypertension, obesity, excessive alcohol; and in later life: social isolation, air pollution and vision loss.

Dementia is an umbrella term encompassing over 200 different types with heterogeneous clinical presentation [5]. This variation, coupled with challenges in identifying individuals at an appropriate stage of disease, contributes to delayed or missed diagnoses. Consequently, a substantial gap exists between the underlying prevalence of dementia and the number of recorded diagnoses. Underdiagnosis has important implications for patient care as a primary care diagnosis is key to accessing the dementia care pathway, enabling referral to specialist memory assessment services and the development of an individualised care plan [6]. Furthermore, an accurate differential diagnosis is essential for appropriate clinical management, as dementia subtypes differ in their underlying pathology, clinical presentation, prognosis, and treatment requirements.

The discrepancy between underlying dementia prevalence and recorded diagnoses is determined by the rate of case ascertainment. Variation in case ascertainment reflects the influence of interacting social and healthcare determinants; said determinants do not operate through independent causal pathways, making it difficult to disentangle their individual contributions [7]. Socioeconomic deprivation represents an important upstream factor, influencing both healthcare access and the prevalence of dementia-related risk factors. However, its relationship with diagnosis rates is complex and contested. Some studies have reported higher diagnosis rates in more deprived areas, potentially reflecting increased general practice contact through the management of comorbid conditions [8], whereas others have identified lower diagnosis rates, attributed to persistent barriers in accessing healthcare [9, 10]. Lower diagnosis rates have also been associated with single-handed general practices [11], which are disproportionately located in rural and deprived areas [12].

Ethnicity represents a further axis of inequality in dementia diagnosis, with individuals from minoritised communities disproportionately affected by dementia [13], and less likely to receive a formal diagnosis [14]. This disparity is likely driven by multiple interacting mechanisms. First, there are broader, non-disease-specific barriers to healthcare access arising from structural and systemic racism. Dementia-specific barriers, including cultural stigma and differing understandings of cognitive decline, may further discourage, or delay, engagement with diagnostic services. Additionally, in some communities, greater reliance on family- and community-based care for older adults may reduce the perceived need to seek a formal diagnosis.

Accurate measurement of diagnosis rates is a longstanding policy concern [15, 16]. Historically the NHS has used financial incentive schemes to encourage GP practices to record dementia diagnoses more systematically, reflecting ongoing concern about under-coding relative to prescribing data. Strategies related to diagnosis, treatment and care are often made at the level of integrated care board (ICB), or sub ICB (SICB), which are the NHS governing bodies of multiple healthcare trusts. Issues with inequitable dementia diagnosis reflect broader concerns about healthcare disparities in the UK [17]. Identifying areas where dementia may be underdiagnosed could provide valuable evidence for informing NHS policy and targeting healthcare resources. Such insights may also support third-sector organisations in directing services more effectively, including the placement of community support services and regional care hubs.

Existing studies of dementia prevalence use NHS estimates [18, 19], which assume case ascertainment rates operate consistently at a given geographic scale. Current NHS and ONS estimates of regional diagnosis use a numerator of observed cases (general practice–level counts of recorded dementia diagnoses), and a denominator of expected prevalence [20] derived from an ageing cohort study adjusted only for age and sex in each region [21]. This approach does not account for potentially important sources of variation, including modifiable risk factors, confounding factors, or spatial effects. Consequently, estimates of expected dementia prevalence derived from this method may be subject to substantial uncertainty.

The increasing availability of large healthcare datasets has enabled the development of more sophisticated statistical approaches for modelling dementia diagnosis. Previous studies have investigated diagnostic gaps, including machine learning approaches to identify potentially undiagnosed individuals from primary care records [22], and data linkage methods to detect underdiagnosis within general practice records [23]. However, no study to date has modelled regional dementia diagnosis rates in England while accounting for the full range of modifiable risk factors.

To address this gap, we develop a multiplicative model of dementia diagnosis rates using openly available public datasets. We define underlying prevalence as the latent proportion of individuals living with dementia, including both diagnosed and undiagnosed cases. The model separates the contribution of underlying disease prevalence from that of healthcare access and quantifies residual geographic variation after accounting for both. We model spatial variation in both underlying dementia prevalence and diagnosis rates, enabling comparison between areas with differing expected diagnostic burden and healthcare capacity to provide differential diagnoses. This analysis complements recent work by Clark et al. [8], which provides a detailed characterisation of barriers to dementia diagnosis in England, and Giebel et al. [9], which examines geographic inequalities in dementia diagnosis and care. By integrating disease prevalence, healthcare access, modifiable risk factors, and differential diagnosis, this study provides a spatial assessment of potential inequalities in dementia diagnosis across England.

## 2 Method

We construct a Bayesian multiplicative model of dementia diagnoses jointly estimating two latent quantities: underlying prevalence which is modelled as a function of risk factors, and the case ascertainment rate which is modelled as a function of healthcare access and sociodemographic barriers. All measures are drawn from publicly available data, at the administrative geography of Local Authority Districts (LADs).

### 2.1 Variables

Figure 1 compares the regional distribution of the population aged 65+ with diagnosed dementia prevalence across England. The spatial patterns suggest that age structure alone does not account for regional variation in diagnosed prevalence, this motivates the multiplicative decomposition into underlying prevalence and case ascertainmentts developed below.

**Figure 1:**
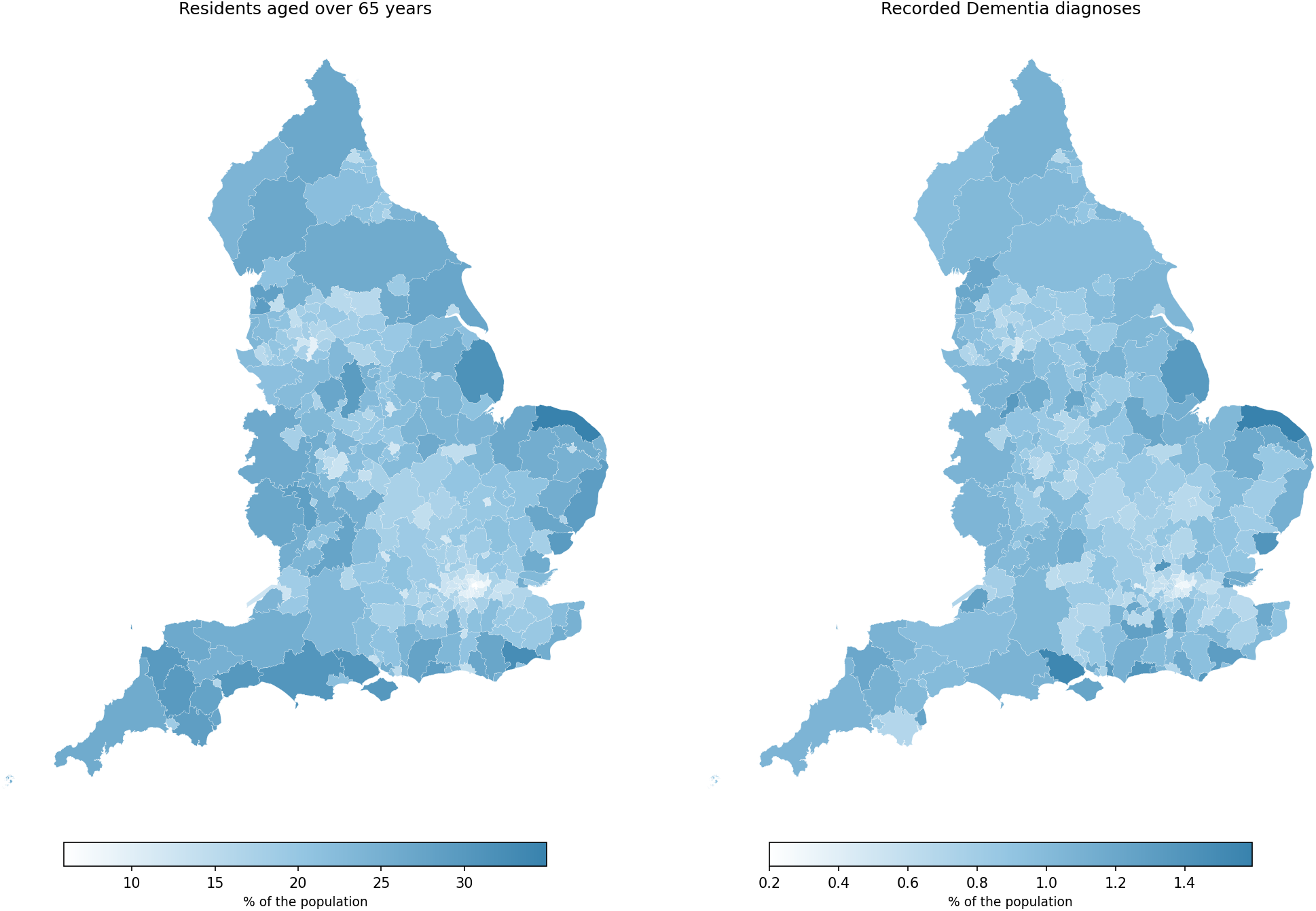
Two maps of England showing the distribution of residents aged over 65 years (left), and the distribution of recorded dementias cases (right).

The dependent variable is observed cases of dementia diagnosis derived from the NHS primary care dementia dataset, as of March 2026. Given the established underdiagnosis of dementia, recorded cases represent a conservative estimate of the underlying disease prevalence. Therefore, we make the simplifying assumption that all recorded dementia diagnoses are correctly classified.

The independent variables are split into two categories: factors affecting underlying prevalence, and factors affecting diagnosis. Table 1 provides further details.

**Table 1:** The factors identified for inclusion in the model, and the corresponding measurements used for them. We were able to find public data for all modifiable risk factors at the correct administrative unit, apart from traumatic brain injury.

| **Category** | **Factor** | **Directly measured (% of LAD population)** | **Proxy measurement (% of LAD population)** |
|---|---|---|---|
| Dependent variable | Dementia | dementia diagnosis |  |
| Non-modifiable risk factor | Age 65+ | over 65 years |  |
| Non-modifiable risk factor | Sex | over 65 years who are female |  |
| Non-modifiable risk factor | Genetics | - | - |
| Modifiable risk factor | Low educational attain-ment | no educational qualifications |  |
| Modifiable risk factor | Hearing loss |  | adults with hearing loss above 25dBHL |
| Modifiable risk factor | High cholesterol | high cholesterol or prescribed statins |  |
| Modifiable risk factor | Depression | adults with depression |  |
| Modifiable risk factor | Traumatic brain injury | - | - |
| Modifiable risk factor | Physical inactivity | adults physically inactive |  |
| Modifiable risk factor | Diabetes | adults with diabetes |  |
| Modifiable risk factor | Smoking | smokers |  |
| Modifiable risk factor | Hypertension | hypertension |  |
| Modifiable risk factor | Obesity | obesity |  |
| Modifiable risk factor | Excessive alcohol |  | alcohol related deaths |
| Modifiable risk factor | Social isolation |  | adults often or always lonely |
| Modifiable risk factor | Air pollution | neighbourhoods with very high pollution (NO2/PM2.5 is double WHO guidelines) |  |
| Modifiable risk factor | Visual loss |  | adults with new sight loss in-cidence |
| Diagnosis factors | GPs per practice | number of GPs (FTE) |  |
| Diagnosis factors | Black ethnicity | Black ethnicity |  |
| Diagnosis factors | Asian ethnicity | Asian ethnicity |  |
| Diagnosis factors | Population density | Population per km<sup>2</sup> |  |
| Diagnosis factors | Economic prosperity |  | adult population claiming unemployment benefit |

#### 2.1.1 Prevalence variables

We split the prevalence variables into non-modifiable and modifiable risk factors, informed by the 2024 Lancet Commission framework on dementia prevention, intervention, and care [4]. We included non-modifiable risk factors of age and sex, both obtained from Office for National Statistics (ONS) mid-year population estimates. Genetic susceptibility, including APOE genotype [2], was not incorporated because aggregated population-level estimates are not publicly available at the geographical scale required for this analysis.

Modifiable risk factors identified by the Lancet Commission were incorporated where appropriate data were available. These included educational attainment; high cholesterol, depression, hypertension, diabetes, obesity, and smoking prevalence; physical inactivity; excessive alcohol consumption (proxied by alcohol-related mortality); social isolation (derived from a social isolation index and household composition measures); air pollution exposure (defined as neighbourhoods exceeding WHO recommended thresholds for PM_2.5_ and PM_10_); and incident vision and hearing loss. Traumatic brain injury was the only modifiable risk factor identified by the Lancet Commission that was excluded, owing to the absence of suitable geographically aggregated data.

#### 2.1.2 Diagnosis variables

Covariates associated with variation in dementia diagnosis rates were selected based on previous literature and conceptual frameworks of healthcare access. These included healthcare provision, ethnicity, socioeconomic deprivation, and population density. Healthcare provision was quantified as the number of full-time equivalent general practitioners (FTE GPs) per LAD, estimated by allocating practice-level workforce data to LADs based on practice catchment area boundaries. Ethnic composition was derived from the 2021 Census, with separate measures for Asian and Black populations. Deprivation was represented using claimant count as a proxy measure of socioeconomic disadvantage, and population density used the mid year population estimates.

#### 2.1.3 Causal structure

Figure 2 demonstrates the assumed relationship between variables, specified using a directed acyclic graph (DAG). We model the diagnosis variables as acting as confounders of underlying prevalence, via the causal pathway of risk factors, and diagnosed cases of dementia.

**Figure 2:**
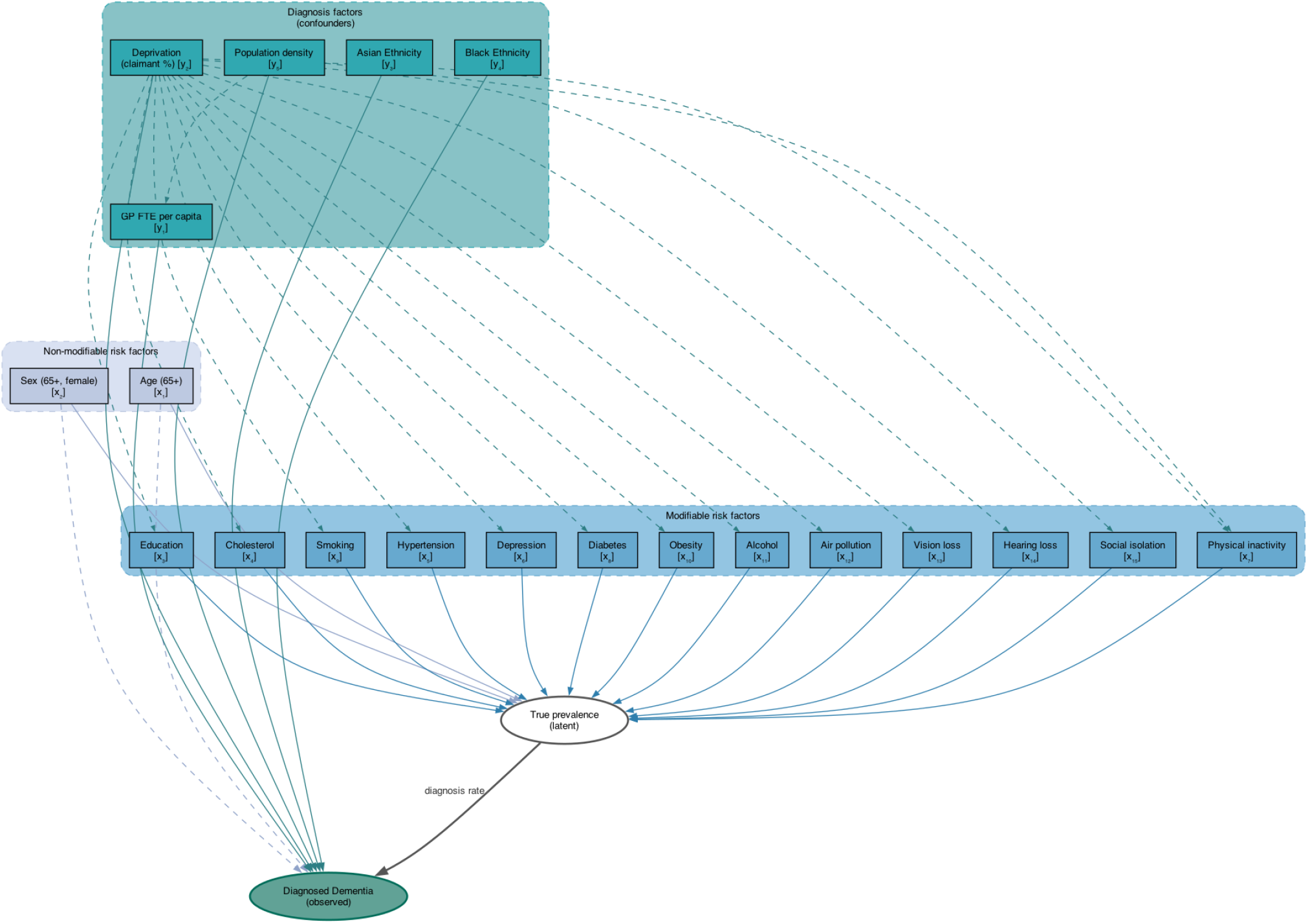
Directed acyclic graph (DAG) illustrating the relationship between variables. Non-modifiable risk factors are taken from the most recent Lancet commission on Dementia [4].

Although the risk factors identified by the Lancet Commission are independently associated with dementia burden [4], these determinants are correlated at an aggregate level. This correlation introduces potential collinearity between predictor variables. To mitigate this, variables capturing similar underlying constructs were combined through mean aggregation.

### 2.2 Data

Data was available at three levels of spatial granularity: GP practices (6,204 in England as of August 2024), LADs (292 in England), and sub-ICBs (106 in England). GP-level data was aggregated to LAD, the primary administrative unit for this analysis. Secondary analysis of differential diagnoses was conducted at sub-ICB level.

As highlighted by The Centre for Public Data [24], significant gaps remain in the availability of routinely collected data on health conditions and NHS staffing at consistent administrative geographies. Hence where necessary spatial joins were used to aggregate the data to a consistent administrative unit; for some variables appropriate proxy measures were used. Full details of factors, measurements, spatial scale, temporal scale, and data sources are detailed in the supplementary data spreadsheet.

Missing values were present for a small number of variables (‘alcohol consumption’, ‘physical inactivity’, ‘hearing loss’, and ‘sight loss’; and were imputed using mean values from the smallest available containing geographic region, further details are given in the Supplementary Material.

All continuous covariates were standardised prior to model fitting to improve parameter estimation and allow model coefficients to represent deviations from the population mean.

### 2.3 Multiplicative Model

We model the expected proportion of the LAD (*i* ∈ *I*) with a recorded diagnosis θ_*i*_ as the product of two unobservable variables: underlying dementia prevalence and case ascertainment. Specifically,

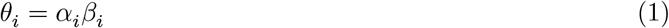

where α_*i*_ is the underlying dementia prevalence and β_*i*_ ∈ [0, 1] represents the probability that an individual with dementia receives a formal diagnosis. Both α and β are estimated as latent variables within a Bayesian framework, with uncertainty propagated through the joint posterior. There is no separate additive error term, as all stochastic variation is captured through the likelihood.

For each LAD, *i* ∈ *I*, underlying dementia prevalence is modelled using a linear predictor of modifiable and non-modifiable risk factors. The predicted prevalence is constrained approximately to the interval ((0,0.02]) through an inverse-logit transformation and scaling factor, corresponding to a maximum prevalence of 40 cases per 1,000 individuals:

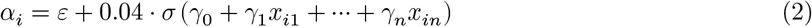

where *x*_*ij*_ represents the *j*th modifiable or non-modifiable risk factor for LAD *i* (as described in Section 2.1.1), σ(·) denotes the inverse-logit function, and ε is a small positive constant introduced to ensure strictly positive prevalence estimates.

For each LAD *i* ∈ *I*, the case ascertainment rate is modelled as a linear predictor of diagnosis related factors:

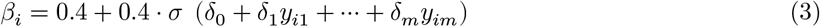

where *y*_*ij*_ represents the *j*th diagnosis-related factor for LAD *i* (Section 2.1.2). This parametrisation constrains diagnosis rates to the interval (0.4, 0.8), reflecting plausible values based on existing evidence. The upper bound encompasses published national estimates for England (approximately 66% [25]), while the lower bound is consistent with lower estimates reported in global meta-analyses [26]. These constraints reduce the range of implausible model solutions.

The observed number of recorded dementia diagnoses *D*_*i*_ in LAD *i* is modelled using a Binomial likelihood:

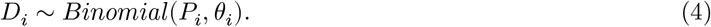

where *P*_*i*_ is the LAD population size.

#### 2.3.1 Constraints

In addition to the constraints placed on α and β via the inverse-logit transformations in Equations 2–3, we also constrain a subset of variable coefficients. The sign constraints were chosen in accordance with the Lancet Commission on Dementia Prevention, Intervention, and Care [27] (see supplementary material); all risk factors with clear individual-level causal evidence were constrained positive (α_*j*_ ≥ 0). Two variables were left unconstrained. First, the proportion of women aged over 65 years was unconstrained since the relationship between sex and dementia prevalence is ambiguous due to variation across dementia subtypes [1]. Secondly, air pollution was unconstrained, since the empirical ecological sign relationship conflicts with the individual-level Lancet evidence due to confounding with urban age structure.

In addition to ensuring the outputs remain within physiologically plausible ranges, these constraints also limit coefficient scale indeterminacy and sign-flip degeneracy which regression models with large numbers of correlated predictors are particularly susceptible to. We refer to this as the sign-constrained model and use it as the basis for all substantive inference.

### 2.4 Spatial Models

To investigate whether geographic clustering remained after accounting for known risk factors, we introduce two spatial models incorporating structured spatial effects.

#### 2.4.1 Multilevel Model (ICB Random Intercepts)

The multilevel model extends the baseline model by incorporating a multilevel structure to account for clustering of diagnostic ascertainment within ICBs. Since ICBs are responsible for commissioning local healthcare services and developing dementia care pathways, an ICB-level random intercept is included in the case ascertainment component β to capture residual variation in diagnosis probability between healthcare systems.

ICBs commission local healthcare services and set dementia diagnosis pathways, to evaluate their influence on the identification of cases ICB-level random intercepts are added to the diagnosis rate β:

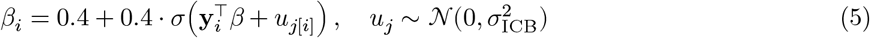

where *j*[*i*] indexes the ICB containing LAD *i* and *u*_*j*_ is an ICB-level random intercept. The variance parameter 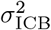 quantifies the extent of between-ICB heterogeneity in case ascertainment after accounting for observed diagnosis-related covariates.

The intraclass correlation coefficient (ICC) estimates the proportion of residual variance in case ascertainment attributable to ICB-level clustering:

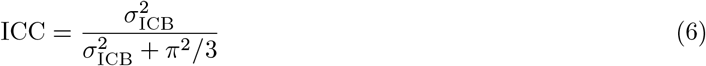

using the standard logistic-normal variance approximation.

#### 2.4.2 Spatial ICAR Model

The second spatial model extends the baseline multiplicative model by incorporating an Intrinsic Conditional Autoregressive (ICAR) spatial random effect into the prevalence component. This captures residual spatial dependence in underlying dementia prevalence between neighbouring LADs after accounting for observed modifiable and non-modifiable risk factors. The inclusion of the ICAR term reflects the assumption that geographically adjacent areas may share unmeasured determinants of disease prevalence, resulting in correlated residual prevalence estimates.

The prevalence component is therefore modelled as:

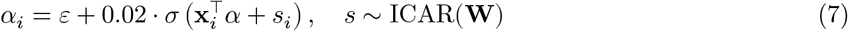

where **W** is a Queen’s contiguity spatial weights matrix and *s*_*i*_ represents the spatially structured random effect for LAD *i*. The magnitude of spatial variation is controlled by the scaling parameter σ_*s*_, estimated from the data with an Exponential(1) prior. Larger posterior estimates of σ_*s*_ indicate greater residual spatial dependence in prevalence after accounting for observed covariates. A soft sum-to-zero constraint 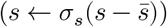 is imposed for identifiability, where 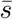 is the mean spatial effect.

Both spatial models use the same sign constraints and prior bounds as the baseline model (Section 2.3.1).

### 2.5 Model fitting

Each model was fitted using Hamiltonian Monte Carlo with the No-U-Turn Sampler (HMC-NUTS) implemented in Stan (via CmdStanPy, [28]), running 4 independent chains in parallel with 1,000 warm-up and 1,000 sampling iterations per chain, yielding 4,000 post-warm-up draws.

### 2.6 Sensitivity Analysis and Evaluation

Multicollinearity among covariates was evaluated using variance inflation factors (VIF). Model convergence was assessed using 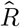 statistics (threshold < 1.01) a and visual inspection of posterior trace plots. Model performance across specifications was compared using root mean squared error (RMSE) calculated against observed recorded dementia diagnoses, alongside the Watanabe–Akaike information criterion (WAIC) to assess Bayesian model fit while accounting for model complexity. Residual spatial dependence was assessed using Moran’s *I* statistic with a permutation test based on 999 permutations. Spatial patterns in model residuals were further examined through choropleth maps of LAD-level residual variation.

Due to potential collinearity of variables we also fitted an unconstrained model with the 13 modifiable prevalence variables reduced using principal component analysis (PCA). This was used as a performance ceiling against which the sign-constrained model was evaluated via RMSE. Further details are available in the Supplementary Material.

## 3 Results

Across the 296 lower-tier local authorities (LADs) in England, the mean recorded dementia prevalence in March 2026 was 8.88 cases per 1,000 population. Prevalence ranged from 1.99 cases per 1,000 in the City of London to 15.95 cases per 1,000 in North Norfolk. As expected, recorded prevalence was strongly correlated with population age structure (*r* = 0.79).

### 3.1 Model performance

Table 2 summarises the performance of the candidate models. The sign-constrained model, in which coefficient directions were informed by prior evidence from the Lancet Commission, achieved a root mean squared error (RMSE) of 0.1199%. To assess the presence of residual spatial structure, two model extensions were considered. Incorporating ICB-level random intercepts produced a modest improvement in model fit (ΔRMSE = 0.0126%, ΔWAIC = 313.1). Extending the prevalence model with an ICAR spatial random effect resulted in the largest improvement (ΔRMSE = 0.1123%, ΔWAIC = 6910.4), indicating substantial residual spatial dependence beyond that explained by the observed covariates.

**Table 2:** Performance of the candidate models. RMSE (root mean squared error) quantifies prediction error between observed and predicted values, while WAIC (Watanabe–Akaike information criterion) evaluates Bayesian model fit while accounting for model complexity. Lower values indicate better model performance.

| Model | Spatial random effect? | RMSE (%) | WAIC |
|---|---|---|---|
| Sign-constrained | no | 0.1199 | 10117.2 |
| Multilevel ICB | yes | 0.1073 | 9804.1 |
| ICAR in prevalence | yes | 0.0076 | 3206.8 |

Given the high degree of collinearity among the explanatory variables, an additional model using principal component analysis (PCA) was evaluated. This yielded only a marginal improvement over the sign-constrained model, suggesting that the informative priors and sign constraints adequately stabilised coefficient estimation despite the presence of multicollinearity.

### 3.2 Sign-constrained model

In the sign-constrained model, all covariates included in the prevalence component were associated with statistically significant coefficients. Age had the largest estimated effect (β = 0.17), consistent with its established role as the strongest population-level risk factor for dementia. The second largest effect was observed for sex, with LADs containing a higher proportion of women exhibiting higher estimated prevalence after adjustment for the remaining covariates.

Among the modifiable risk factors, all variables except air pollution were positively associated with dementia prevalence. Air pollution was the only covariate with a negative coefficient, reflecting its inverse association with prevalence after adjustment for the remaining variables. This is likely attributable to its correlation with more densely populated urban areas, which generally have younger population structures.

The largest positive coefficients among the modifiable risk factors were observed for incident hearing loss (+0.045), alcohol-related mortality (+0.044), lower educational attainment (+0.021), relative to: depression, incident vision loss, social isolation, cardiovascular comorbidities, unhealthy lifestyle, and physical inactivity (all ≤ 0.001).

In the sign-constrained model all diagnosis-related covariates had posterior distributions that excluded zero. Population density had the largest positive coefficient (+0.7460). Greater socioeconomic deprivation (proxied by claimant count) and higher proportions of Asian and Black populations were associated with lower case ascertainment, whereas FTE GP availability had a positive, albeit comparatively small, association.

Figure 3 presents the posterior coefficient estimates for the prevalence and diagnosis covariates across the three model specifications. Differences in coefficient magnitudes across models are an expected consequence of the scale indeterminacy inherent in the joint estimation of two latent variables.

**Figure 3:**
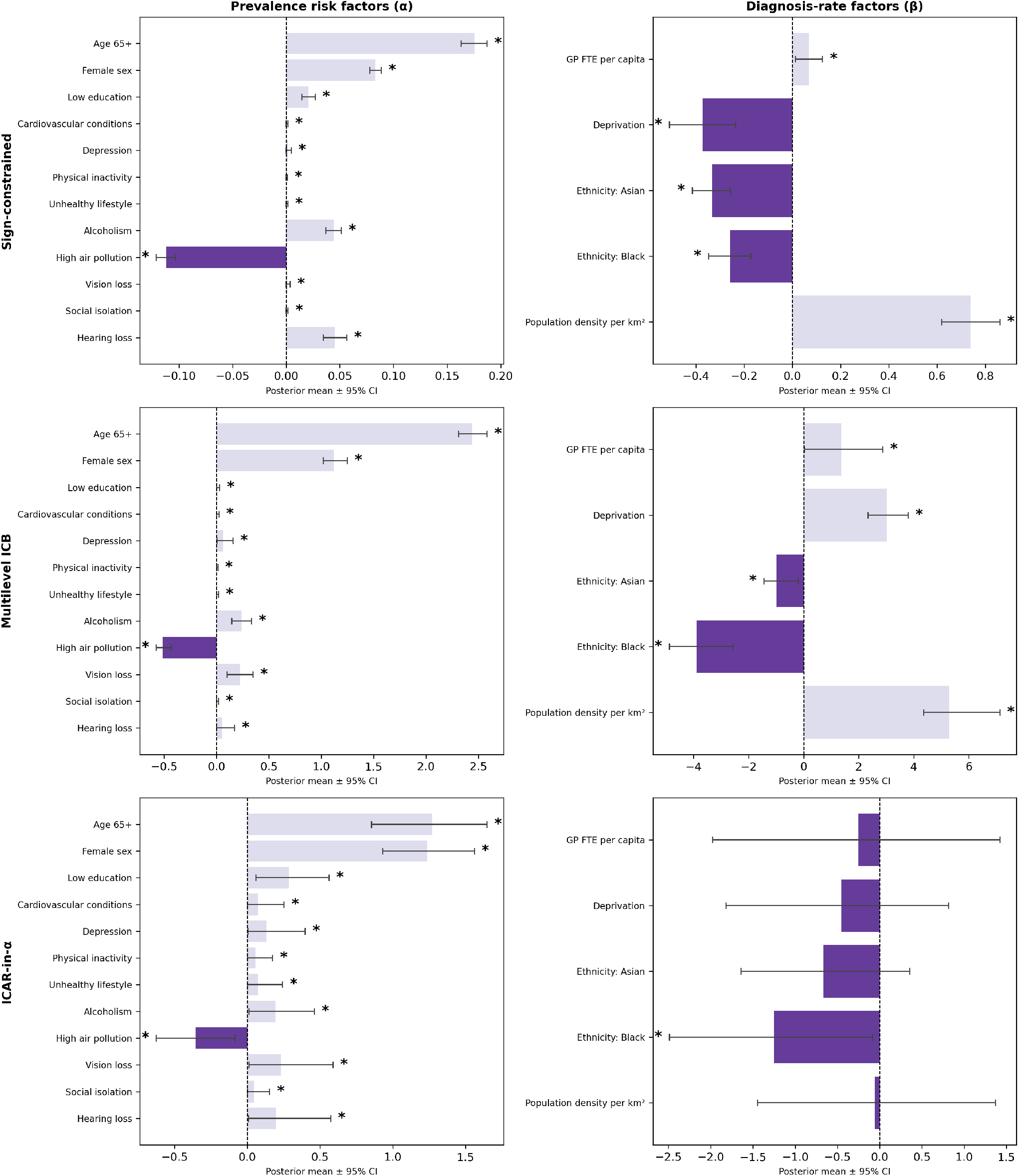
The coefficients for the variables in the baseline sign constrained model, the multilevel ICB model, and the spatial random effect in prevalence (ICAR-in-α) model. Coefficients for the underlying prevalence factors are shown left, and case ascertainment factors right. The bars show the 95% confidence interval, with * indicating that the posterior confidence interval excludes 0.

### 3.3 Spatial effects

As stated, both the multilevel ICB model, and spatial ICAR model improved on the sign-constrained model. For the multilevel ICB model the intraclass correlation coefficient was 0.341(95%*CI* : 0.203–0.597), indicating that 34% of the residual variation in the case ascertainment rate β logit was attributable to ICB membership, pointing to the importance of commissioning decisions and local healthcare infrastructure.

Figure 4 compares estimated true prevalence (left) and case-ascertainment rate (right) across the three primary models. Adding a spatial random effect to prevalence (ICAR-in-α) absorbs more residual spatial variation, pushing fitted prevalence down and diagnosis rate up relative to the non-spatial baseline. Since only the product (α × β) is identified by the observed data of dementia cases, this compensating shift reflects that the two components cannot be fully separated, and their absolute scales depend partly on where spatial structure is placed in the model.

**Figure 4:**
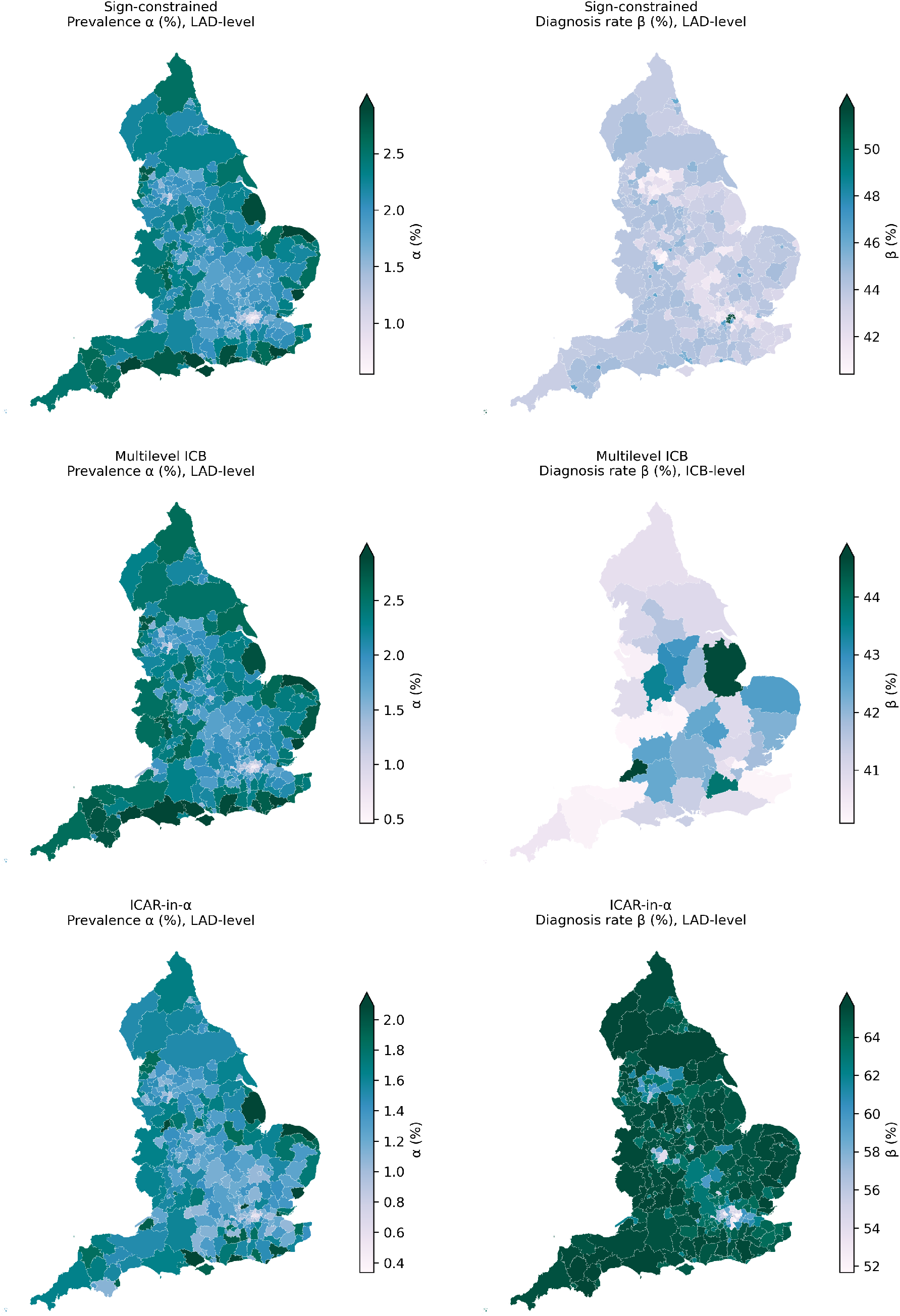
Choropleth maps showing the estimated underlying prevalence (left) and estimated case ascertainment rate (right) for the three models.

### 3.4 Differential diagnosis

Across England, Alzheimer’s disease accounted for an average of 40.00% of recorded dementia diagnoses, followed by vascular dementia (13.39%), mixed dementia (8.32%), dementia with Lewy bodies (2.96%), and frontotemporal dementia (0.60%). A further 0.73% of diagnoses were assigned to less common dementia subtypes not explicitly represented in the NHS coding framework. However, 24.18% of diagnoses were recorded as unspecified dementia, with an additional 1.82% classified as inconclusive. Considerable geographic variation was observed in the recording of dementia subtypes across sub-ICBs, as demonstrated by Figure 5.

**Figure 5:**
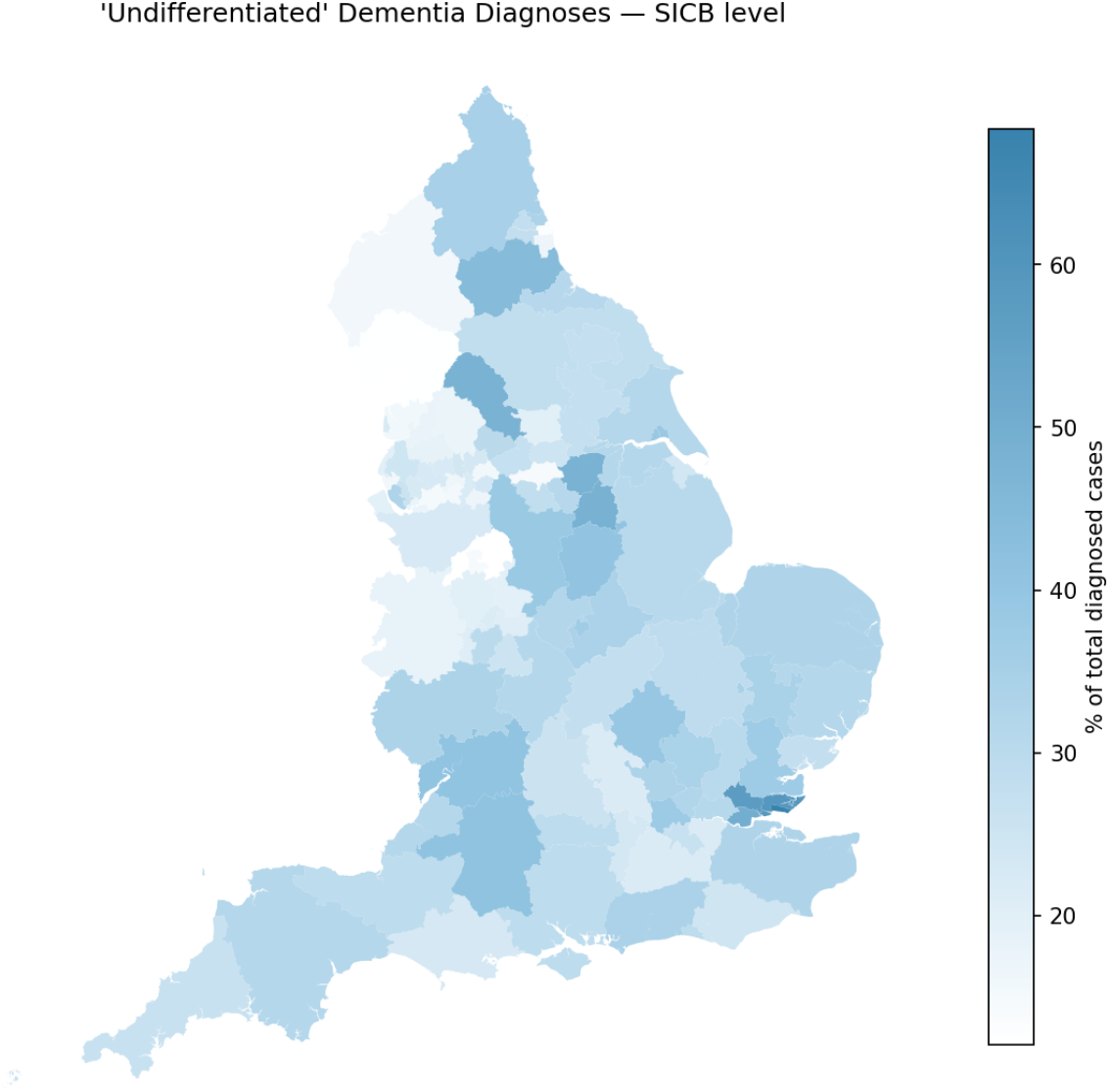
Choropleth map demonstrating the variation in the percent of undifferentiated dementia diagnoses at sub-ICB level. If all individuals received the recommended level of dementia care, the number of undifferentiated cases should be negligible.

Figure 6 compares estimated under diagnosis with the proportion of unspecified dementia diagnoses. Areas shown in dark blue exhibit both lower estimated case ascertainment and higher proportions of unspecified dementia diagnoses.

**Figure 6:**
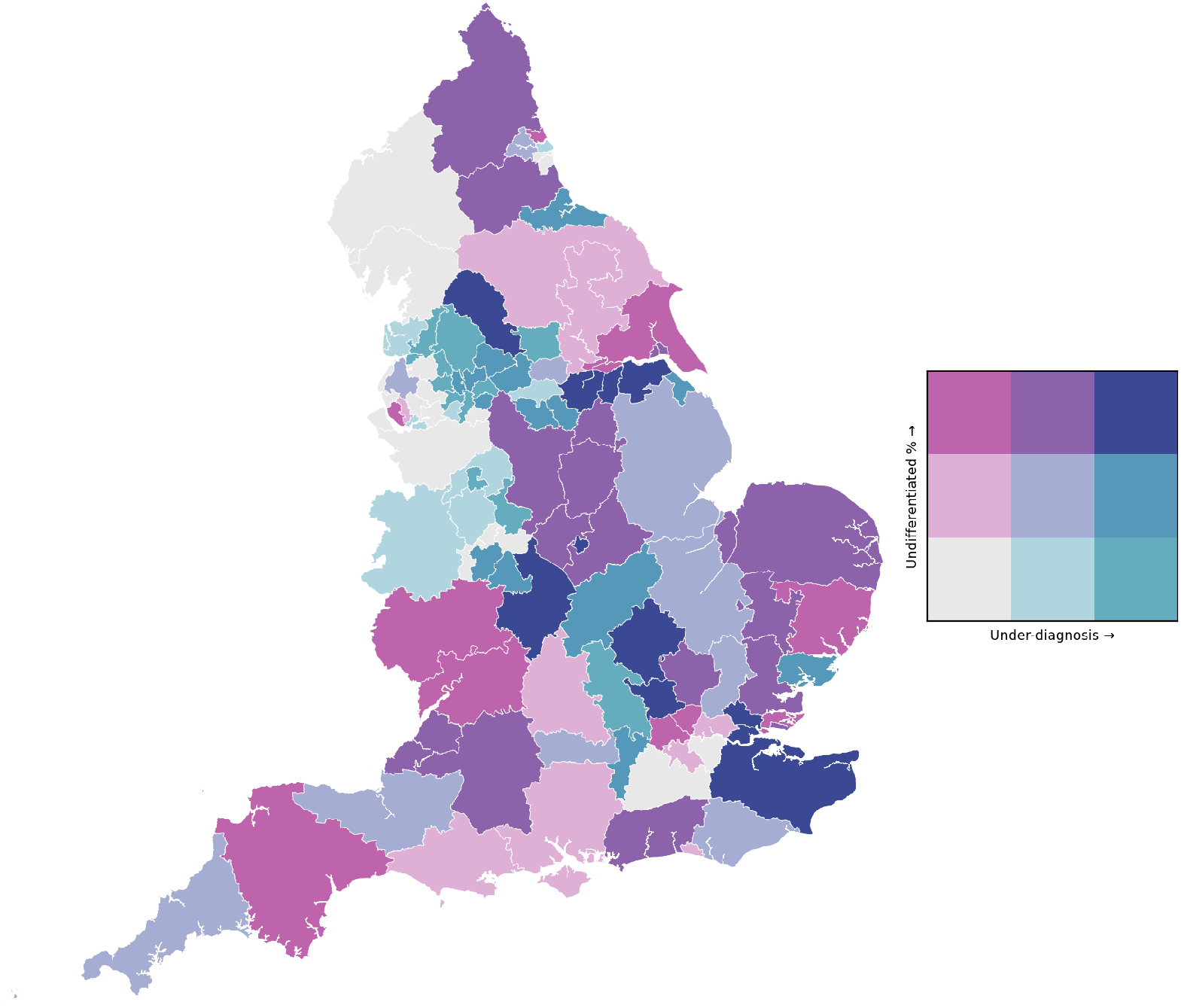
Bivariate choropleth map showing the relationship between model-estimated underdiagnosis (from the sign-constrained baseline model) and the proportion of undifferentiated dementia diagnoses at sub-ICB level. Colour categories are relative across regions; all areas exhibit some degree of incomplete case ascertainment and incomplete differential diagnosis. Regions in dark blue represent areas with comparatively high levels of both underdiagnosis and undifferentiated dementia coding, and may therefore represent priorities for further investigation and targeted improvement of diagnostic pathways.

## 4 Discussion

This work jointly estimates underlying dementia prevalence and case ascertainment using routinely collected administrative data, with the goal of understanding regional variation in diagnosis. By distinguishing latent disease prevalence from case ascertainment, our approach enables a more nuanced analysis than previous studies, which have often used recorded diagnoses as a proxy for prevalence [18, 19]. This distinction helps disentangle regional variation in underlying disease prevalence from differences in regional access to diagnosis. By estimating posterior distributions for all model parameters, the framework quantifies the magnitude and uncertainty of each non-modifiable and modifiable risk factor, as well as factors affecting diagnosis access, enabling assessment of their relative importance. Through combined modelling with rates of unspecified diagnoses, we identify regions where targeted care resources could have the highest impact.

The underlying prevalence model identified age as the dominant predictor followed by sex according with the established literature on risk factors for dementia [4], and hence supporting the epidemiological validity of the modelling framework. Among modifiable risk factors, incident hearing loss showed the strongest association, consistent with evidence identifying hearing loss as both a major risk factor and a potential early marker of neurodegeneration [29, 30]. In contrast, air pollution showed an unexpected negative association with latent dementia prevalence, contrary to the literature [31, 32]. A possible explanation is the influence of spatial differences in case ascertainment, confounding the fact that air pollution is strongly correlated with urbanicity. Although our model accounted for population density, the use of aggregate LAD-level data may limit our ability to disentangle the effects of pollution on underlying disease burden from geographical variation in diagnosis. In particular, higher case ascertainment in urban areas may offset the group level increase in dementia burden associated with air pollution.

The model identified substantial inequalities in dementia case ascertainment, of which population density was the strongest predictor. Higher ascertainment in urban areas is consistent with established rural–urban disparities in access to healthcare [33, 34]. Socioeconomic deprivation was also consistently associated with lower ascertainment, reflecting the role of deprivation as an upstream determinant influencing both modifiable dementia risk factors across life course, and access to healthcare. This reinforces the need for dementia prevention and diagnostic strategies that address wider social determinants of health [17].

Ethnicity was significantly associated with case ascertainment, with areas containing larger Asian and Black populations exhibiting lower diagnosis rates. This finding is consistent with evidence of barriers to dementia diagnosis within minoritised communities, including stigma, differences in healthcare engagement, and greater reliance on informal care networks [14, 35]. These populations have historically been underserved [36], and require greater community engagement to improve access to diagnosis. The consistently larger negative coefficient for Asian ethnicity highlights the heterogeneity in diagnostic inequalities across ethnic minority groups, reinforcing the need for outreach and service provision to be tailored to the needs of specific communities rather than treating ethnic minorities as a homogeneous population [37].

Through the inclusion of the number of GPs, and the multilevel modelling of ICBs, we aimed to quantify the impact of NHS service provision on case ascertainment, to understand the ‘postcode lottery of care’. Although a higher number of GPs, and therefore potentially greater availability of primary care, was positively associated with case ascertainment,the magnitude of this association was smaller than we had anticipated [11]. One possible explanation relates to the highly right-skewed distribution of GPs across LADs (see supplementary material), with a small number of areas having substantially higher per-capita GP workforce levels than others. It is plausible that these areas have already been targeted for enhanced healthcare investment, but that any downstream improvement in diagnosis rates has yet to be captured in administrative statistics, a lag that would be expected given the time required for workforce expansion to translate into clinical outcomes. In contrast to the relatively small coefficient for GP count, the multilevel model demonstrated that 34% of residual variation in case ascertainment was attributable to ICB membership. Given that ICBs commission local healthcare services and dementia pathways [6], this finding suggests that regional organisation and delivery of care represent important determinants of diagnostic equity.

The best-fitting model incorporated spatial correlation in underlying dementia prevalence between neighbouring areas, suggesting that regional variation in dementia rates may partly reflect unmeasured determinants of disease risk not fully captured by the model covariates. Further research is needed to identify these factors, which could include differences in particulate matter exposure not captured by the air pollution variable included in our model, which previous studies have linked to dementia [38, 39]. Access to more granular data would enable a more detailed investigation of geographic heterogeneity in both underlying prevalence, and diagnosis rates.

Beyond overall diagnostic coverage, the analysis identified substantial geographic variation in diagnostic specificity. A staggeringly large proportion of dementia diagnoses were recorded without a specified subtype, with marked differences observed between sub-ICBs. This accords with findings of high correlation between deprivation (measured using the index of multiple deprivation) and unspecified dementia diagnoses [10]. This is clinically significant, as dementia subtypes often differ in their underlying pathology, prognosis, and management [40], and accurate differential diagnosis is becoming increasingly important with the emergence of disease-specific therapies [41]. By combining estimates of low case ascertainment with rates of unspecified diagnoses, the model identifies regions where targeted improvements to diagnostic services could have the greatest impact, including Yorkshire, Warwickshire, Central East, Essex, and Kent. We interpret these findings in the context of a model that assumes case ascertainment remains below 100% in all areas, emphasising that improvements to diagnostic services are needed nationally.

Several limitations should be considered. Firstly, although the model incorporates all modifiable risk factors identified by the Lancet Commission [27], their interpretation at the area level requires caution. Aggregate LAD-level measures capture population distributions of risk rather than individual exposure histories and therefore cannot establish individual causal pathways [42]. Furthermore, prevalence and case ascertainment are latent quantities, jointly estimated from the same observed diagnosis data, meaning that estimates of α and β in the model should be interpreted as geographic patterns rather than definitive causal effects. Secondly, there is the caveat of data availability and quality; several variables were represented using proxy measures, which may introduce unmodelled variation as they do not fully capture the underlying risk factors. Since the analysis relied on public data, there is also uncertainty from differences in data collection methods and temporal alignment between sources.

An important limitation is the need to contextualise diagnosis rates within broader debates about what constitutes a ‘good’ dementia diagnosis. An ideal diagnosis should be both timely and accurate [6]. However, because dementia symptoms may emerge years after the underlying biological processes begin, there is an inevitable lag between disease onset and diagnosis, as is the case for many long-term conditions [43]. Consequently, a snapshot of recorded diagnoses cannot fully capture the temporal dimensions of diagnostic access, including whether individuals are diagnosed promptly following symptom onset. More granular longitudinal data would enable future research to examine these temporal dynamics and provide a more nuanced understanding of variation in quality of dementia diagnosis across areas.

### 4.1 Further Work

Beyond the risk factors identified by the Lancet commission, there could be further characteristics affecting underlying disease prevalence. This analysis captures the most commonly reported barriers to diagnosis; however, a recent report by Clark et al. [8] identifies disability and sexuality as additional factors associated with inequitable access to diagnosis. The inclusion of these variables could be explored in future work.

Beyond diagnosis rates, further geographic analysis could examine inequalities in access to post-diagnostic care and support. The quality of dementia care is shaped in part by the availability of, and access to, health, social care and third-sector services [6]. Although our model accounts for variation in GP provision and population density, it does not directly capture geographical accessibility to healthcare providers or downstream support services. This highlights two important avenues for further research. First, spatial analysis could examine how easily people can physically access diagnostic and post-diagnostic services, accounting for factors such as travel distances and service availability [44]. Second, research could investigate the relationship between case ascertainment and access to post-diagnostic care, including health and social care services offered through charities and community groups. Together, these analyses could help establish whether geographical inequalities in diagnosis are accompanied by additional inequalities in access to ongoing care and support, and identify where these intersecting gaps are greatest. This motivates the development of a systematic atlas of dementia care service availability in England, which would allow the geographic patterns identified here to be linked to care access outcomes. This would allow modelling of both availability and accessibility using gravity models [45] and enable more targeted guidance on the location of care services relevant to forthcoming NHS transformations in health and social care service delivery [46].

## 5 Conclusion

This study presents a scalable Bayesian framework that separates underlying dementia prevalence from case ascertainment, providing a more nuanced understanding of geographic inequalities in diagnosis than analyses based on recorded diagnosis rates alone. Our findings demonstrate substantial geographic variation in dementia diagnosis across England, persisting after accounting for non-modifiable and modifiable risk factors. By identifying areas with low case ascertainment and poor diagnostic specificity, the framework provides an evidence base for more targeted and equitable dementia policy and service planning. The substantial contribution of ICB-level variation further highlights the importance of locally commissioned healthcare pathways in addressing diagnostic inequalities. Ultimately, improving equity in dementia care requires not only increasing diagnosis rates, but also ensuring timely and accurate differential diagnosis to facilitate personalised care and appropriate treatment pathways.

## Data Availability

The study only uses public data - URL links to all the public datasets are provided in the supplementary material.

## Acknowledgements

BT is supported by UKRI EPSRC Grant No. EP/Y028392/1: AI for Collective Intelligence (AI4CI). The Dementia Research Centre is supported by the Alzheimer’s Society, Alzheimer’s Research UK and The Wolfson Foundation. JJ has been supported by a Frontotemporal Dementia Research Studentship in Memory of David Blechner (funded through The National Brain Appeal) and NIHR (NIHR306146). JDW acknowledges funding support from Alzheimer’s Research UK, the Alzheimer’s Society, the Royal National Institute for Deaf People and the UCLH NIHR Biomedical Research Centre. SJC acknowledges funding support from Marie Curie and Alzheimer’s Society. The views expressed in this publication are those of the authors and not necessarily those of the NIHR or the UK Department of Health and Social Care.

## Notes

### Competing Interest Statement

The authors have declared no competing interest.

## References

1. Podcasy JL and Epperson CN. Considering sex and gender in Alzheimer disease and other dementias. Dialogues in clinical neuroscience 2016; 18:437–46. doi: 10.31887/DCNS.2016.18.4/cepperson

2. Williams DM, Heikkinen S, Hiltunen M, Davies NM, and Anderson EL. The proportion of Alzheimer’s disease attributable to apolipoprotein E. npj Dementia 2026 Jan; 2:1–. doi: 10.1038/s44400-025-00045-9

3. Corrada MM, Brookmeyer R, Paganini-Hill A, Berlau D, and Kawas CH. Dementia Incidence Continues to Increase with Age in the Oldest Old The 90+ Study. Annals of Neurology 2010; 67:114. doi: 10.1002/ANA.21915

4. Livingston G, Huntley J, Liu KY, Costafreda SG, Selbæk G, Alladi S, Ames D, Banerjee S, Burns A, Brayne C, Fox NC, Ferri CP, Gitlin LN, Howard R, Kales HC, Kivimäki M, Larson EB, Nakasujja N, Rockwood K, Samus Q, Shirai K, Singh-Manoux A, Schneider LS, Walsh S, Yao Y, Sommerlad A, and Mukadam N. Dementia prevention, intervention, and care: 2024 report of the Lancet standing Commission. The Lancet 2024 Aug; 404:572–628. doi: 10.1016/S0140-6736(24)01296-0

5. Prince M, Knapp M, Geurhcet M, Mccrone P, and Prina M. Dementia UK: update. Tech. rep. Alzheimer’s Society, 2014

6. National Collaborating Centre for Mental Health. The Dementia Care Pathway: Full Implementation Guidance. Tech. rep. Updated October 2018. London: National Collaborating Centre for Mental Health, 2018. Available from: https://www.rcpsych.ac.uk/docs/default-source/improving-care/nccmh/dementia/nccmh-dementia-care-pathway-full-implementation-guidance.pdf

7. Walker IF, Lord PA, and Farragher TM. Variations in dementia diagnosis in England and association with general practice characteristics. Primary Health Care Research & Development 2017; 18:235–41. doi: 10.1017/S146342361700007X

8. Clark M, Compton L, Salehi N, Knapp M, and Comas-Herrera A. Research on dementia and inequalities in care and support – a map of the terrain and discussion of key issues. International Journal for Equity in Health 2026 Dec; 25:82. doi: 10.1186/S12939-025-02601-4

9. Giebel C, Readman MR, Godfrey A, Gray A, Carton J, and Polden M. Geographical inequalities in dementia diagnosis and care: A systematic review. International Psychogeriatrics 2025 Jun; 37:100051. doi: 10.1016/J.INPSYC.2025.100051

10. Jitlal M, Amirthalingam GN, Karania T, Parry E, Neligan A, Dobson R, Noyce AJ, Marshall CR, and Kostev K. The Influence of Socioeconomic Deprivation on Dementia Mortality, Age at Death, and Quality of Diagnosis: A Nationwide Death Records Study in England and Wales 2001-2017. Journal of Alzheimer’s Disease 2021; 81:321–8. doi: 10.3233/JAD-210089

11. Connolly A, Gaehl E, Martin H, Morris J, and Purandare N. Underdiagnosis of dementia in primary care: variations in the observed prevalence and comparisons to the expected prevalence. Aging & mental health 2011; 15:978–84. doi: 10.1080/13607863.2011.596805

12. Szymczynska P, Innes A, Mason A, and Stark C. A Review of Diagnostic Process and Postdiagnostic Support for People With Dementia in Rural Areas. Journal of Primary Care & Community Health 2011; 2:262–76. doi: 10.1177/2150131911404705

13. Mukadam N, Marston L, Lewis G, Mathur R, Rait G, and Livingston G. Incidence, age at diagnosis and survival with dementia across ethnic groups in England: a longitudinal study using electronic health records. Alzheimer’s & Dementia 2023; 19:1300–7

14. Subramaniam H and Ladinska EBM. Systemic disadvantages facing UK ethnic elders within dementia healthcare. BJPsych Advances 2025 Sep; 31:311–20. doi: 10.1192/BJA.2025.10115

15. Alzheimer’s Society. Increasing access to a dementia diagnosis. https://www.alzheimers.org.uk/what-we-do/policy-and-influencing/increasing-access-dementia-diagnosis. Accessed: 2026-10-19. 2024 May

16. All-Party Parliamentary Group on Dementia. Fuelling the Moonshot: Unleashing the UK’s potential through dementia research. Tech. rep. Accessed: 2026-10-19. London, 2021 Sep. Available from: https://www.alzheimers.org.uk/sites/default/files/2021-09/Fuelling_Moonshot_APPG.pdf

17. Marmot M. Health equity in England: the Marmot review 10 years on. BMJ 2020 Feb; 368. doi: 10.1136/BMJ.M693

18. Knapp M, Prince M, Albanese E, Banerjee S, Dhanasiri S, Fernandez JL, Ferri C, Snell T, and Stewart R. Dementia UK: The full report. Tech. rep. Alzheimer’s Society, 2007

19. Ferri CP, Prince M, Brayne C, Brodaty H, Fratiglioni L, Ganguli M, Hall K, Hasegawa K, Hendrie H, Huang Y, Jorm A, Mathers C, Menezes PR, Rimmer E, and Scazufca M. Global prevalence of dementia: A Delphi consensus study. The Lancet 2005 Dec; 366:2112–7. doi: 10.1016/S0140-6736(05)67889-0

20. NHS England Digital. Recorded Dementia Diagnoses: Methodology of Indicators. Last edited 11 July 2022. Accessed 28 October 2026. 2022. Available from: https://digital.nhs.uk/data-and-information/publications/statistical/recorded-dementia-diagnoses/recorded-dementia-diagnoses-supporting-information/recorded-dementia-diagnoses-methodology

21. Matthews FE, Arthur A, Barnes LE, Bond J, Jagger C, Robinson L, and Brayne C. A two-decade comparison of prevalence of dementia in individuals aged 65 years and older from three geographical areas of England: Results of the cognitive function and ageing study i and II. The Lancet 2013 Oct; 382:1405–12. doi: 10.1016/S0140-6736(13)61570-6

22. Jammeh EA, Carroll CB, Pearson Stephen W, Escudero J, Anastasiou A, Zhao P, Chenore T, Zajicek J, and Ifeachor E. Machine-learning based identification of undiagnosed dementia in primary care: a feasibility study. BJGP Open 2018 Jul; 2. doi: 10.3399/BJGPOPEN18X101589

23. Aldus CF, Arthur A, Dennington-Price A, Millac P, Richmond P, Dening T, Fox C, Matthews FE, Robinson L, Stephan BC, Brayne C, and Savva GM. Journals Library Undiagnosed dementia in primary care: a record linkage study. Health Services and Delivery Research 2020; 8. doi: 10.3310/hsdr08200

24. Centre for Public Data. Data Gaps in Healthcare Statistics. Tech. rep. March 2024. Centre for Public Data, 2024 Mar. Available from: https://static1.squarespace.com/static/5ee7a7d964aeed7e5c507900/t/660568dcb7722e3a819c03ef/1711630577258/gaps-in-healthcare-statistics.pdf

25. NHS England. Dementia Diagnoses in England at Record High. https://www.england.nhs.uk/2024/07/dementia-diagnoses-in-england-at-record-high/. Published 22 July 2024. 2024 Jul

26. Lang L, Clifford A, Wei L, Zhang D, Leung D, Augustine G, Danat IM, Zhou W, Copeland JR, Anstey KJ, and Chen R. Prevalence and determinants of undetected dementia in the community: a systematic literature review and a meta-analysis. BMJ Open 2017 Feb; 7:e011146. doi: 10.1136/BMJOPEN-2016-011146

27. Chen Y, Bandosz P, Stoye G, Liu Y, Wu Y, Lobanov-Rostovsky S, French E, Kivimaki M, Livingston G, Liao J, and Brunner EJ. Dementia incidence trend in England and Wales, 2002–19, and projection for dementia burden to 2040: analysis of data from the English Longitudinal Study of Ageing. The Lancet Public Health 2023 Nov; 8:e859–e867. doi: 10.1016/S2468-2667(23)00214-1

28. Stan Development Team. Stan Reference Manual, V2.39. 2026

29. Lin FR, Metter EJ, O’Brien RJ, Resnick SM, Zonderman AB, and Ferrucci L. Hearing Loss and Incident Dementia. Archives of Neurology 2011 Feb; 68:214–20. doi: 10.1001/ARCHNEUROL.2010.362

30. Johnson JC, Marshall CR, Weil RS, Bamiou DE, Hardy CJ, and Warren JD. Hearing and dementia: from ears to brain. Brain 2021 Mar; 144:391–401. doi: 10.1093/BRAIN/AWAA429

31. Peters R, Ee N, Peters J, Booth A, Mudway I, and Anstey KJ. Air Pollution and Dementia: A Systematic Review. Journal of Alzheimer’s Disease 2019 Feb; 70:S145–S163. doi: 10.3233/JAD-180631

32. Chen H, Kwong JC, Copes R, Hystad P, Donkelaar A van, Tu K, Brook JR, Goldberg MS, Martin RV, Murray BJ, et al. Exposure to ambient air pollution and the incidence of dementia: a population-based cohort study. Environment international 2017; 108:271–7. doi: 10.1016/j.envint.2017.08.020

33. Todd A, Copeland A, Husband A, Kasim A, and Bambra C. Access all areas? An area-level analysis of accessibility to general practice and community pharmacy services in England by urbanity and social deprivation. BMJ Open 2015 May; 5:e007328. doi: 10.1136/BMJOPEN-2014-007328

34. Deaney C, Hemingway E, Reesby D, and Scott V. Evaluating the Care Gaps and Inequalities Arising in Continuing Healthcare (CHC) Delivery in Rural Areas. Journal of Primary Care and Community Health 2024 Jan; 15. doi: 10.1177/21501319241305968

35. Mukadam N, Cooper C, Basit B, and Livingston G. Why do ethnic elders present later to UK dementia services? A qualitative study. International Psychogeriatrics 2011 Sep; 23:1070–7. doi: 10.1017/S1041610211000214

36. Krishnamurthy S and Rollin FG. We must be clear that the root cause of racial disparities in Alzheimer’s disease is racism. Alzheimer’s & Dementia 2023; 19:5305–6. doi: 10.1002/alz.13389

37. Webster L, Amador S, Rapaport P, Mukadam N, Sommerlad A, James T, Javed S, Roche M, Lord K, Bharadia T, et al. Tailoring STrAtegies for RelaTives for Black and South Asian dementia family carers in the United Kingdom: A mixed methods study. International Journal of Geriatric Psychiatry 2023; 38:e5868. doi: 10.1002/gps.5868

38. Russ TC, Gatz M, Pedersen NL, Hannah J, Wyper G, Batty GD, Deary IJ, and Starr JM. Geographical variation in dementia: examining the role of environmental factors in Sweden and Scotland. Epidemiology (Cambridge, Mass.) 2015; 26:263. doi: 10.1097/EDE.0000000000000230

39. Jones A, Ali MU, Mayhew A, Aryal K, Correia RH, Dash D, Manis DR, Rehman A, O’Connell ME, Taler V, et al. Environmental risk factors for all-cause dementia, Alzheimer’s disease dementia, vascular dementia, and mild cognitive impairment: An umbrella review and meta-analysis. Environmental research 2025; 270:121007. doi: 10.1016/j.envres.2025.121007

40. Taylor B, Mohamud S, Brotherhood E, Harding E, Waddington C, Camic PM, Alexander D, Crutch S, Stott J, Hardy C, et al. Decline in activities of daily living in the rarer dementias. General Psychiatry 2025; 38:e101905. doi: 10.1136/gpsych-2024-101905

41. Giebel C, Silva-Ribeiro W, Watson J, Volkmer A, Chirico I, Diaz A, Heath B, Hanna K, and Talbot C. A systematic review on the evidence of misdiagnosis in dementia and its impact on accessing dementia care. International Journal of Geriatric Psychiatry 2024; 39:e6158. doi: 10.1002/gps.6158DigitalObjectIdentifier(DOI)

42. Leyland AH and Groenewegen PP. Multilevel modelling and public health policy. Scandinavian journal of public health 2003; 31:267–74. doi: 10.1080/14034940210165028

43. Anosova O, Head A, Collins B, Alexiou A, Darras K, Sutton M, Cookson R, Anselmi L, O’Flaherty M, Barr B, et al. Estimating the burden of underdiagnosis within England: a modelling study of linked primary care data. PLoS One 2025; 20:e0313877. doi: 10.1371/journal.pone.0313877

44. Todd A, Copeland A, Husband A, Kasim A, and Bambra C. Access all areas? An area-level analysis of accessibility to general practice and community pharmacy services in England by urbanity and social deprivation. BMJ open 2015; 5:e007328. doi: 10.1136/bmjopen-2014-007328

45. Stacherl B and Sauzet O. Gravity models for potential spatial healthcare access measurement: a systematic methodological review. International Journal of Health Geographics 2023 Dec; 22:34–. doi: 10.1186/S12942-023-00358-Z

46. Department of Health and Social Care. Fit for the Future: 10 Year Health Plan for England. Tech. rep. CP 1350. London: Department of Health and Social Care, 2025 Jul. Available from: https://www.gov.uk/government/publications/10-year-health-plan-for-england-fit-for-the-future

