## Supplementary_tables_figures for "Mapping inequalities in dementia diagnosis in England: a Bayesian spatial analysis of disease prevalence and case ascertainment"

November 9, 2026

#### Method

##### Data

The modelling framework used exclusively public data. As demonstrated in Figure 1, there was data missing ness for Alcohol deaths, physical inactivity, hearing loss, and sight loss. As the data was available at multiple administrative units, missing data entries were filled with data for the smallest containing region with a valid value.

Figure 2 presents the unadjusted distributions of all model variables. As expected, relatively stable demographic variables, such as age and sex, and measures of general health exhibited approximately normal distributions, whereas more heterogeneous factors, including pollution and claimant count, showed substantial skew. Prior to modelling, all variables were standardised using z-scores, allowing coefficients to represent variation relative to the population mean.

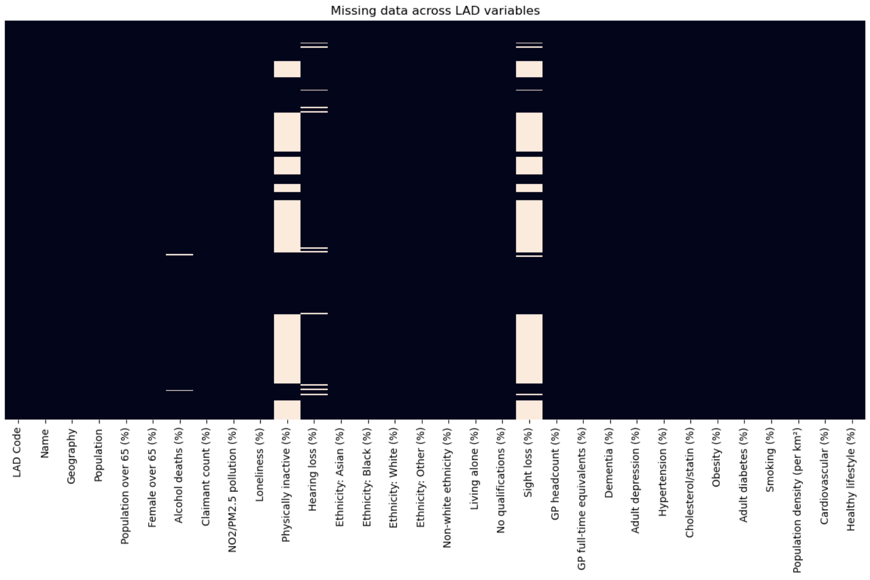


Figure 1: Plot showing where data was missing, it was filled with data for the smallest containing geographic area.

Table 1 reports the unadjusted correlations between the candidate prevalence variables and recorded dementia cases per 1,000 population. The sign constraints applied in the model were informed by the *Lancet* Commission on dementia. No sign constraint was imposed on sex because its association with dementia prevalence at the

Table 1: The raw correlation between the variables and dementia, and the sign-constraints imposed in the baseline sign-constrained model.

| Variable | Correlation with dementia | Sign-constraint |
|---|---|---|
| Age | +0.603 | +1 |
| Sex | +0.072 | - |
| Educational attainment | -0.017 | +1 |
| Cardiovascular | -0.059 | +1 |
| Depression | +0.036 | +1 |
| Physically inactive | -0.027 | +1 |
| Unhealthy lifestyle | -0.028 | +1 |
| Alcoholism | +0.068 | +1 |
| Air pollution | -0.061 | - |
| Visual loss | -0.025 | +1 |
| Loneliness | -0.002 | +1 |
| Hearing loss | -0.009 | +1 |

population level remains uncertain. Likewise, no sign constraint was applied to air pollution, as its aggregate spatial relationship with dementia is difficult to disentangle from the confounding effects of urban areas.

#### PCA model

As stated in the main text, due to potential collinearity of variables we also fitted a model with variables reduced using principal component analysis (PCA). PCA was applied to the 13 modifiable prevalence variables, reducing them to a smaller set of four orthogonal components (Figure 3). The resulting components, combined with age and sex, were used to fit an unconstrained version of the multiplicative model. This was used as a predictive benchmark, providing a performance ceiling against which the sign-constrained model was evaluated via root mean squared error.

Figure 4 demonstrates the model results, with the coefficients for the PCA components shown. The PCA model achieved $RMSE = 0.1192\%$ and $WAIC = 19016.6$, demonstrating a modest improvement on the sign-constrained model in RMSE $\Delta RMSE = 0.0007x$. Whilst this model was a marginal improvement on the sign constrained model, due to the lack of interpretability the sign constrained model was preferentially used.

### Results

Figure 5 is a scatter plot showing the quartile ranges for the sub-ICBs across the domains of diagnosis rate and diagnosis quality.

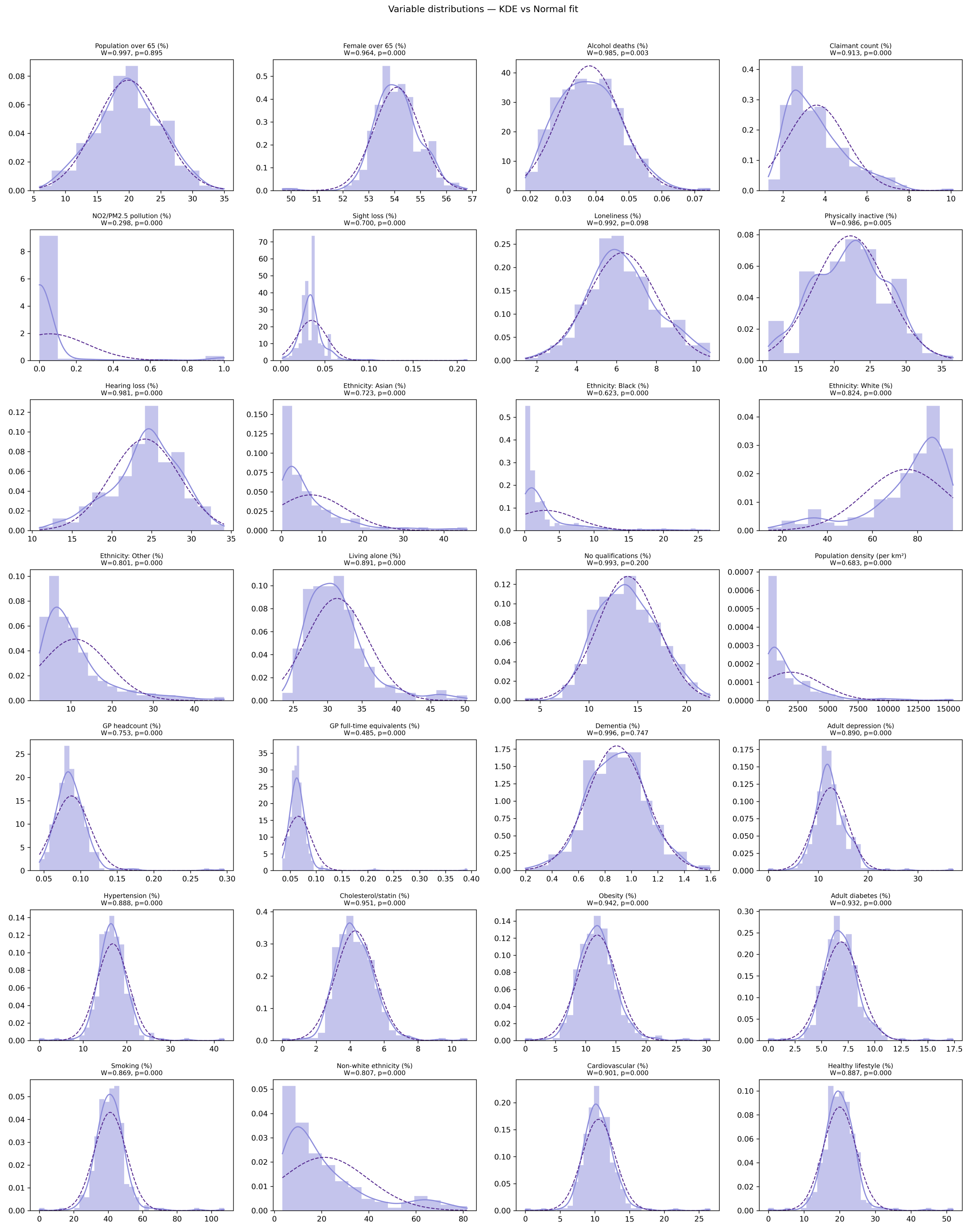


Figure 2: Histograms for all the variables included in the model.

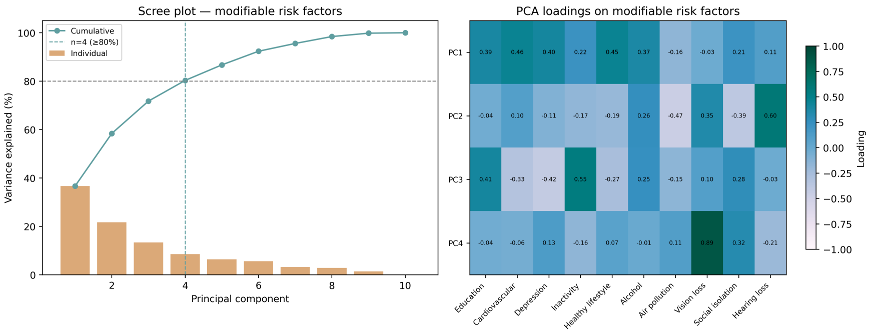


Figure 3: Plot showing the PCA loadings.

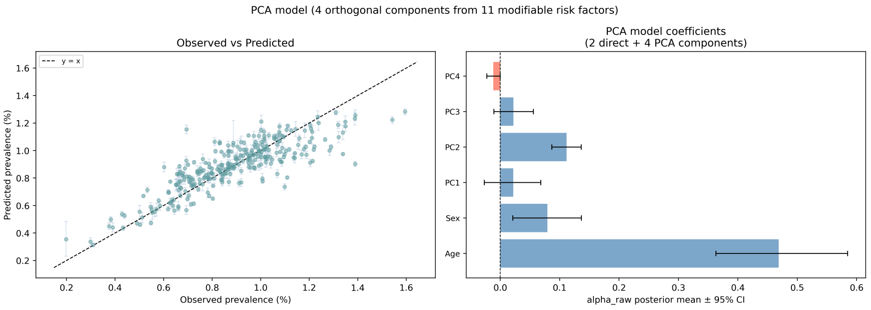


Figure 4: Plot showing the PCA model results.

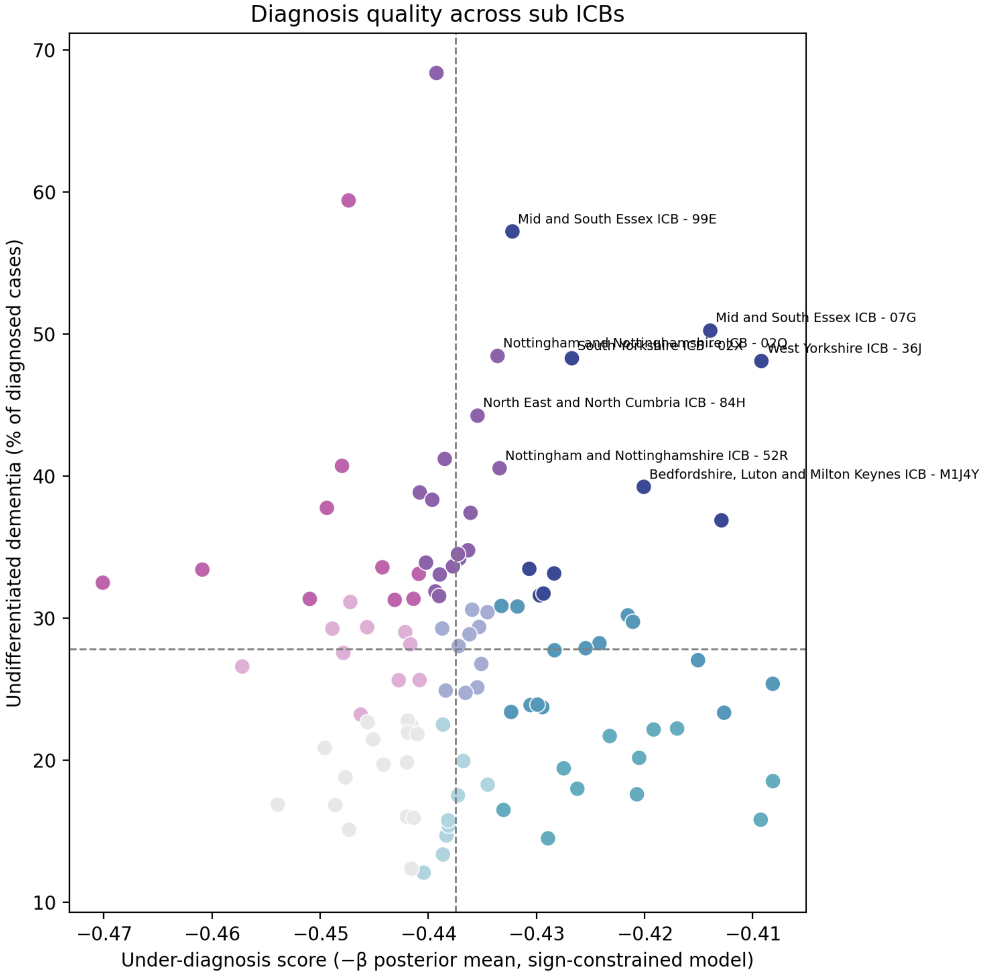


Figure 5: Scatter diagram showing the sub ICBs with lowest model predicted rates.
